# Missing Outcomes and Treatment Effect Estimates in the A4 Study of Solanezumab

**DOI:** 10.64898/2026.11.20.26368608

**Authors:** Eileen Y. Jang, Kendra D. Sims, Michael Flanders, Sarah F. Ackley

## Abstract

**INTRODUCTION:** Missing final-visit outcome measurements may bias treatment effect estimates in amyloid-targeting drug trials. We examined whether accounting for missing Week-240 outcomes changed estimated treatment differences in the Anti-Amyloid Treatment in Asymptomatic Alzheimer’s Disease (A4) trial.

**METHODS:** We reanalyzed 1,169 randomized participants, assessing baseline-to-Week-240 change in amyloid positron emission tomography (PET) Centiloids, the Preclinical Alzheimer Cognitive Composite (PACC), and Clinical Dementia Rating–Sum of Boxes (CDR-SB). We compared available-case estimates with inverse probability of censoring weighting (IPCW; primary analysis) and multiple imputation under missing at random (MAR), and assessed departures from MAR with delta-adjusted missing not at random (MNAR) analyses.

**RESULTS:** Week-240 amyloid PET, PACC, and CDR-SB were missing for 352 (30.1%), 345 (29.5%), and 343 (29.3%) participants. IPCW treatment differences (solanezumab minus placebo) were -8.74 Centiloids (95% bootstrap CI, -11.75 to -5.86), -0.076 PACC points (CI, -0.691 to 0.517), and 0.126 CDR-SB points (CI, -0.077 to 0.320). These were close to the available-case differences of -8.56 Centiloids, -0.100 PACC points, and 0.120 CDR-SB points, and the MAR imputation differences. Assuming missing solanezumab outcomes were 0.50 SD worse than MAR predictions, differences were -3.48 Centiloids (95% CI, -6.50 to -0.46), -0.981 PACC points (CI, -1.602 to -0.360), and 0.297 CDR-SB points (CI, 0.112 to 0.481).

**DISCUSSION:** Methods assuming that missingness is explained by measured data did not change the A4 findings. The amyloid result withstood large departures from this assumption, but PACC and CDR-SB results became unfavorable under modest ones, so conclusions for these outcomes depend on why data are missing.

## Introduction

In trials of amyloid-targeting treatments for Alzheimer’s disease, participants who miss the final visit may differ from those who complete follow-up, so analyses limited to completers may not represent the full randomized population. This is especially important for amyloid positron emission tomography (PET), often measured only at baseline and study end, where a missed final scan precludes estimating change from baseline.

The Anti-Amyloid Treatment in Asymptomatic Alzheimer’s Disease (A4) trial evaluated solanezumab, an anti-amyloid monoclonal antibody, in cognitively unimpaired older adults with elevated brain amyloid. Solanezumab did not slow cognitive decline, although amyloid accumulation was lower than in the placebo group.^1^ A subsequent reanalysis examined whether amyloid change could serve as a surrogate for cognitive decline.^2^ However, in the A4 data package, 352 of 1,169 randomized participants (30.1%) lacked the Week-240 amyloid PET measurement, and 345 (29.5%) and 343 (29.3%) lacked Week-240 Preclinical Alzheimer Cognitive Composite (PACC) and Clinical Dementia Rating-Sum of Boxes (CDR-SB) assessments, respectively. No studies have assessed how missing end-of-study outcomes might affect A4 treatment-effect estimates.

In this study, we reanalyzed all 1,169 randomized A4 participants to assess whether accounting for missing Week-240 outcomes changed the estimated effects of solanezumab on amyloid, cognitive, and functional outcomes. We compared available-case estimates with inverse probability of censoring weighting (IPCW) as our primary approach, as well as with multiple imputation (MI) under missing at random (MAR), and conducted sensitivity analyses under missing not at random (MNAR) assumptions.

## Research Design and Methods

### A4 Study and Study Population

The A4 study was a Phase 3 randomized controlled trial of solanezumab, an anti-amyloid monoclonal antibody, to reduce cognitive decline in individuals with elevated amyloid assessed via amyloid positron emission tomography (PET) but no clinical symptoms.^3^ Additional details are reported elsewhere.^3–6^ All participants provided informed consent at enrollment.^2^ Institutional Review Board (IRB) approval was obtained at each of the trial sites.^2^ Data were obtained from the A4 and Longitudinal Evaluation of Amyloid Risk and Neurodegeneration (LEARN) Study Data Package.^2^ For this study, we included all 1,169 participants randomized in the A4 study and analyzed them according to randomized assignment.

### Amyloid-PET and Magnetic Resonance Imaging

Amyloid-PET imaging was planned for all participants at baseline and Week 240. Standardized uptake value ratios (SUVRs) were converted to Centiloids using the following formula: Centiloids = 183.07 x (Florbetapir SUVR) - 177.26.^2^ Magnetic resonance imaging (MRI) was performed at baseline and at Week 240, and processing is described in detail elsewhere.^7^ We used baseline (pre-randomization) intracranial, forebrain parenchymal, hippocampal, entorhinal, and lateral ventricular volumes, hippocampal occupancy, and microbleed reads as predictors in the missing-outcome models.

### Cognitive and Functional Outcomes

The A4 trial’s primary endpoint was change in the PACC at 4.5 years.^1^ In this study, the primary outcomes were change from baseline to Week 240 in amyloid PET (Centiloids) and the PACC. CDR-SB, with higher scores indicating worse cognitive and functional status, was a secondary outcome measure. For each outcome, change was defined as the Week-240 value minus the baseline value, and treatment differences as solanezumab minus placebo. We refer to PACC and CDR-SB together as cognitive and functional outcomes.

### Assessment of Missing Outcomes

We determined the number of participants without Week-240 measures separately for amyloid PET, PACC, and CDR-SB, overall and by randomized treatment, and compared missingness between arms using chi-square tests. We also reviewed treatment-period disposition record categories among participants with missing Week-240 outcomes.

These categories record why participants left the treatment period and do not necessarily explain why a particular measurement was missing.

### Covariates and Data Preparation

For plasma p-tau217 results outside the limits of quantification, we used recorded raw instrument values; glial fibrillary acidic protein (GFAP) and neurofilament light (NfL) measurements below the quantification limit or with insufficient sample, non-numeric, or duplicate Aβ42/40 results were set to missing. MRI volumes coded as invalid (-4) were set to missing. Microbleeds were taken from the latest pre-randomization MRI read.

Week-240 amyloid PET values were limited to analyzed scans; early-termination scans were not used. Visits and missing values for each variable are listed in Supplementary Table S2.

### Inverse Probability of Censoring Weighting

We first calculated unadjusted mean-change differences among participants with an available Week-240 endpoint (available-case analysis), with heteroskedasticity-robust (HC0) standard errors. For primary IPCW analyses, we modeled the probability of observing each endpoint using logistic regression with treatment assignment, age, sex, APOE-ε4 copy number, baseline amyloid, and baseline PACC. Participants with observed endpoints received inverse observation-probability weights stabilized by the observation rate in their treatment arm. We estimated percentile confidence intervals from 500 bootstrap samples drawn within treatment arms, refitting the observation model in each sample, and examined predicted observation probabilities, maximum weights, and effective sample sizes.

Additional observation models included baseline demographic characteristics, microbleeds, MRI volumes, blood biomarkers, and other cognitive measures such as the Mini-Mental State Exam (MMSE), logical memory delayed recall, and Free and Cued Selective Reminding Test (FCSRT), or all baseline predictors. Missing baseline predictors were filled with median values, recomputed within each bootstrap sample. Primary analyses used untrimmed stabilized weights; as a sensitivity analysis, weights were truncated at their 1st and 99th percentiles. ARIA-E before Week 240 was rare (two definite cases, both in the placebo arm, among 1,144 participants with follow-up MRI).

New microbleeds or superficial siderosis (ARIA-H) were more common (99 [17.0%] placebo vs 84 [14.9%] solanezumab) but, as post-randomization events, were not included in the observation models.

### Multiple Imputation under Missing At Random

We performed multiple imputation by chained equations (MICE) with 50 imputations and 60 iterations. The core imputation model included treatment assignment, age, sex, APOE-ε4 copy number,and the baseline and Week-240 values of all three outcomes.

An extended imputation model also included education, body mass index, depressive symptoms (Geriatric Depression Scale >5), baseline hippocampal volume and presence of microbleeds, plasma Aβ42/40 ratio and p-tau217, baseline MMSE and Digit Symbol Substitution Test (DSST), and Week-96 PACC. Continuous variables were standardized before imputation and restored to their original scales afterward. We used predictive mean matching (PMM)for continuous variables and logistic regression to estimate the probability for binary variables. In each imputed dataset, change from baseline was regressed on treatment assignment. Estimates were combined using Rubin’s rules, with confidence intervals based on a t-distribution with Barnard-Rubin small-sample degrees of freedom. As in the available-case and IPCW analyses, the analysis model included no covariates, so all three approaches estimate the same unadjusted difference in mean change. We assessed trace plots of imputed means and standard deviations, logged events, fractions of missing information, and Monte Carlo error (Supplementary Figure S2).

### Missing Not at Random Sensitivity Analyses

Observed covariates may not capture all reasons for missingness. For missing not at random (MNAR) analyses, we assessed the impact on treatment effects for several scenarios in which participants with missing Week-240 outcomes had lower cognition (lower PACC or higher CDR-SB) or greater amyloid accumulation than predicted under MAR. Starting from the core MAR imputations, we added a deterministic shift to each originally missing value, so each participant’s value was shifted relative to their own MAR-imputed value. Observed outcomes were unchanged. Shifts were based on the standard deviation (SD) of the observed Week-240 endpoint in the placebo arm, and only originally missing outcomes were shifted. We examined six scenarios: (1) no shifts, which reproduces the core MAR analysis; (2) 0.25 SD in both arms; (3) 0.50 SD in both arms; (4) 0.25 SD in the solanezumab arm only; (5) 0.5 SD in the solanezumab arm only; and (6) 0.25 SD in the placebo arm combined with 0.5 SD in the solanezumab arm. In a tipping-point analysis, we increased the shift for missing solanezumab outcomes from 0 to 1 SD in steps of 0.05, with no shift in the placebo arm, and identified the smallest shift at which the 95% CI changed position relative to zero.^8^

These CIs are conditional on the assumed shifts and were not adjusted for multiple comparisons. Analyses were performed in R (version 4.5.2) using the mice package (version 3.19.0).

## Results

### Participant Characteristics and Outcome Availability

Participants had a mean age of 71.9 years (SD, 4.8); 59.4% were female, 94.1% were White, 96.2% were not Hispanic or Latino, and 58.9% carried at least one APOE-ε4 allele. Mean baseline amyloid was 66.1 Centiloids (SD, 32.8). Baseline characteristics were generally similar between groups, although microbleeds were more common in placebo than solanezumab participants (25.1% vs 19.7%; Table 1).

**Table 1:** Baseline characteristics by randomized treatment assignment

| **Characteristic** | **Solanezumab (n = 578)** | **Placebo (n = 591)** | **Overall (n = 1169)** |
|---|---|---|---|
| **Demographics** |  |  |  |
| **Age, years** | 71.9 (4.6) | 71.9 (5.0) | 71.9 (4.8) |
| **Female** | 337 (58.3%) | 357 (60.4%) | 694 (59.4%) |
| **Education, years** | 16.6 (2.7) | 16.5 (2.9) | 16.6 (2.8) |
| **Race** |  |  |  |
| White | 545 (94.3%) | 555 (93.9%) | 1100 (94.1%) |
| Black or African American | 12 (2.1%) | 16 (2.7%) | 28 (2.4%) |
| Asian | 11 (1.9%) | 13 (2.2%) | 24 (2.1%) |
| American Indian or Alaska Native | 1 (0.2%) | 1 (0.2%) | 2 (0.2%) |
| More than one race | 5 (0.9%) | 3 (0.5%) | 8 (0.7%) |
| Unknown or not reported | 4 (0.7%) | 3 (0.5%) | 7 (0.6%) |
| **Ethnicity** |  |  |  |
| Not Hispanic or Latino | 556 (96.2%) | 568 (96.1%) | 1124 (96.2%) |
| Hispanic or Latino | 16 (2.8%) | 18 (3.0%) | 34 (2.9%) |
| Unknown or Not Reported | 6 (1.0%) | 5 (0.8%) | 11 (0.9%) |
| **Clinical and Biomarker** |  |  |  |
| **Amyloid PET, Centiloids** | 66.1 (33.5) | 66.0 (32.1) | 66.1 (32.8) |
| **Geriatric Depression Scale > 5** | 13 (2.2%) | 14 (2.4%) | 27 (2.3%) |
| **BMI, kg/m2** | 27.2 (5.0) | 27.6 (5.2) | 27.4 (5.1) |
| **APOE ε4 copies** |  |  |  |
| 0 | 236 (40.8%) | 244 (41.3%) | 480 (41.1%) |
| 1 | 295 (51.0%) | 300 (50.8%) | 595 (50.9%) |
| 2 | 47 (8.1%) | 47 (8.0%) | 94 (8.0%) |
| **Baseline Cognition** |  |  |  |
| PACC | -0.009 (2.757) | 0.009 (2.604) | -0.000 (2.680) |
| CDR-SB | 0.064 (0.170) | 0.048 (0.164) | 0.056 (0.167) |
| MMSE | 28.8 (1.3) | 28.8 (1.2) | 28.8 (1.3) |
| DSST | 48.5 (10.3) | 48.8 (9.8) | 48.6 (10.0) |
| **Baseline MRI** |  |  |  |
| Intracranial volume, cm<sup>3</sup> | 1476.4 (141.7) | 1470.0 (146.2) | 1473.1 (144.0) |
| Forebrain parenchyma volume, cm<sup>3</sup> | 917.8 (91.8) | 914.7 (95.1) | 916.2 (93.4) |
| Hippocampal volume (left + right), cm<sup>3</sup> | 5.8 (0.7) | 5.8 (0.7) | 5.8 (0.7) |
| Entorhinal volume (left + right), cm<sup>3</sup> | 5.1 (0.8) | 5.1 (0.8) | 5.1 (0.8) |
| Lateral ventricle volume (left + right), cm<sup>3</sup> | 36.4 (17.1) | 36.3 (18.3) | 36.4 (17.7) |
| Hippocampal occupancy | 0.737 (0.086) | 0.736 (0.092) | 0.737 (0.089) |
| Any microbleeds | 114 (19.7%) | 148 (25.1%) | 262 (22.4%) |
| **Baseline Plasma Biomarker** |  |  |  |
| p-tau217, U/mL | 0.286 (0.170) | 0.267 (0.146) | 0.277 (0.159) |
| Aβ42/40 ratio | 0.108 (0.082) | 0.108 (0.079) | 0.108 (0.081) |
| GFAP, ng/mL | 0.122 (0.058) | 0.124 (0.063) | 0.123 (0.061) |
| NfL, pg/mL | 3.4 (1.4) | 3.4 (1.8) | 3.4 (1.6) |

Note: Continuous variables are mean (SD); categorical variables are n (%) among participants with observed values for each variable. ‘Unknown or not reported’ is a recorded race/ethnicity category.
Abbreviations: Aβ, amyloid-beta; APOE, apolipoprotein E; BMI, body mass index; CDR-SB, Clinical Dementia Rating–Sum of Boxes; DSST, Digit Symbol Substitution Test; GFAP, glial fibrillary acidic protein; MMSE, Mini-Mental State Examination; NfL, neurofilament light chain; PACC, Preclinical Alzheimer Cognitive Composite; p-tau217, phosphorylated tau 217.

At Week 240, amyloid PET was available for 817 participants, the PACC for 824, and CDR-SB for 826. The corresponding proportions without these endpoints were 30.1%, 29.5%, and 29.3%. Missingness was slightly more common with solanezumab than placebo for amyloid (31.3% vs 28.9%), PACC (30.6% vs 28.4%), and CDR-SB (29.9% vs 28.8%), but differences between treatment groups were not statistically significant (chi-square tests, all p > 0.3). Among 345 participants missing the Week-240 PACC composite, the most common recorded treatment-period disposition categories were withdrawal by subject (189, 54.8%) and adverse event (64, 18.6%); 11 (3.2%) had death recorded, and nine (2.6%) were lost to follow-up.

### Treatment Effects After IPCW

For amyloid, the treatment difference was -8.56 Centiloids (95% CI, -11.48 to -5.63) in the available-case analysis, and -8.74 (95% bootstrap CI, -11.75 to -5.86) with IPCW (Table 2). For PACC, the difference changed from -0.100 points (95% CI, -0.678 to 0.479) in the available-case analysis to -0.076 (95% bootstrap CI, -0.691 to 0.517) with IPCW. For CDR-SB, the available-case and IPCW differences were 0.120 (95% CI, -0.060 to 0.300) and 0.126 points (95% bootstrap CI, -0.077 to 0.320), respectively. Differences varied little across the seven untrimmed IPCW models (amyloid -8.80 to -8.60; PACC -0.094 to - 0.072; CDR-SB 0.126 to 0.133; Figure 1). Weights were stable (minimum fitted observation probability, 0.33; maximum weight, 1.68), and truncating them changed differences by at most 0.04 Centiloids, 0.004 PACC points, and 0.001 CDR-SB points.

**Figure 1:**
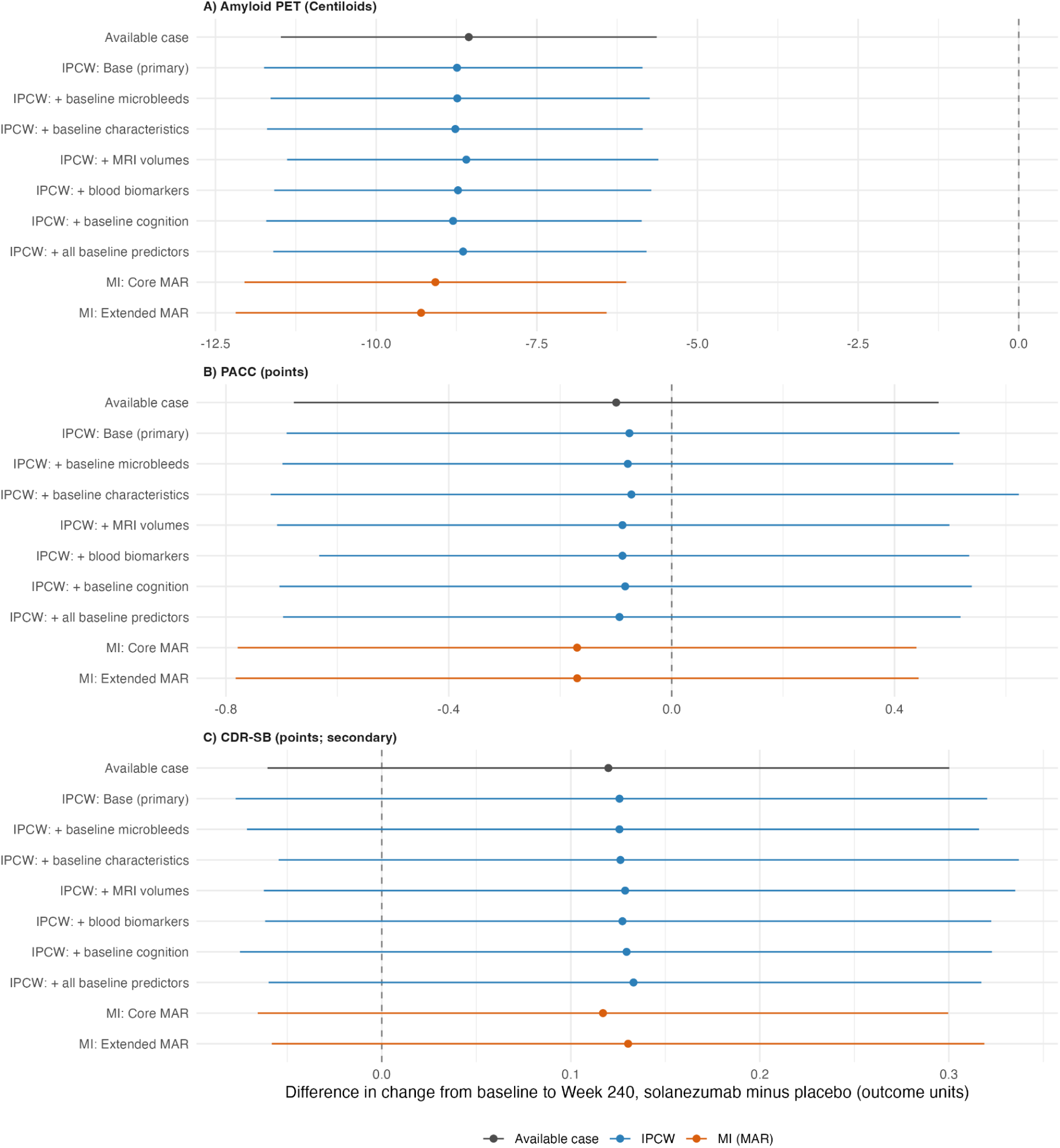
Treatment-effect estimates across missing-outcome analyses in A4. Panel A shows amyloid PET; Panel B shows PACC; Panel C shows CDR-SB (secondary) in adjacent plots. Estimates are standardized by the observed placebo-group SD of baseline-to-Week-240 change; points show differences and bars indicate 95% confidence intervals. IPCW specifications include the primary baseline observation model and versions with additional baseline microbleeds, demographic characteristics, MRI volumes, blood biomarkers, cognitive measures, or all baseline predictors. The available-case and MAR MI estimates are shown for comparison. Negative differences indicate less amyloid accumulation or worse PACC performance, but better CDR-SB outcomes.

**Table 2.**
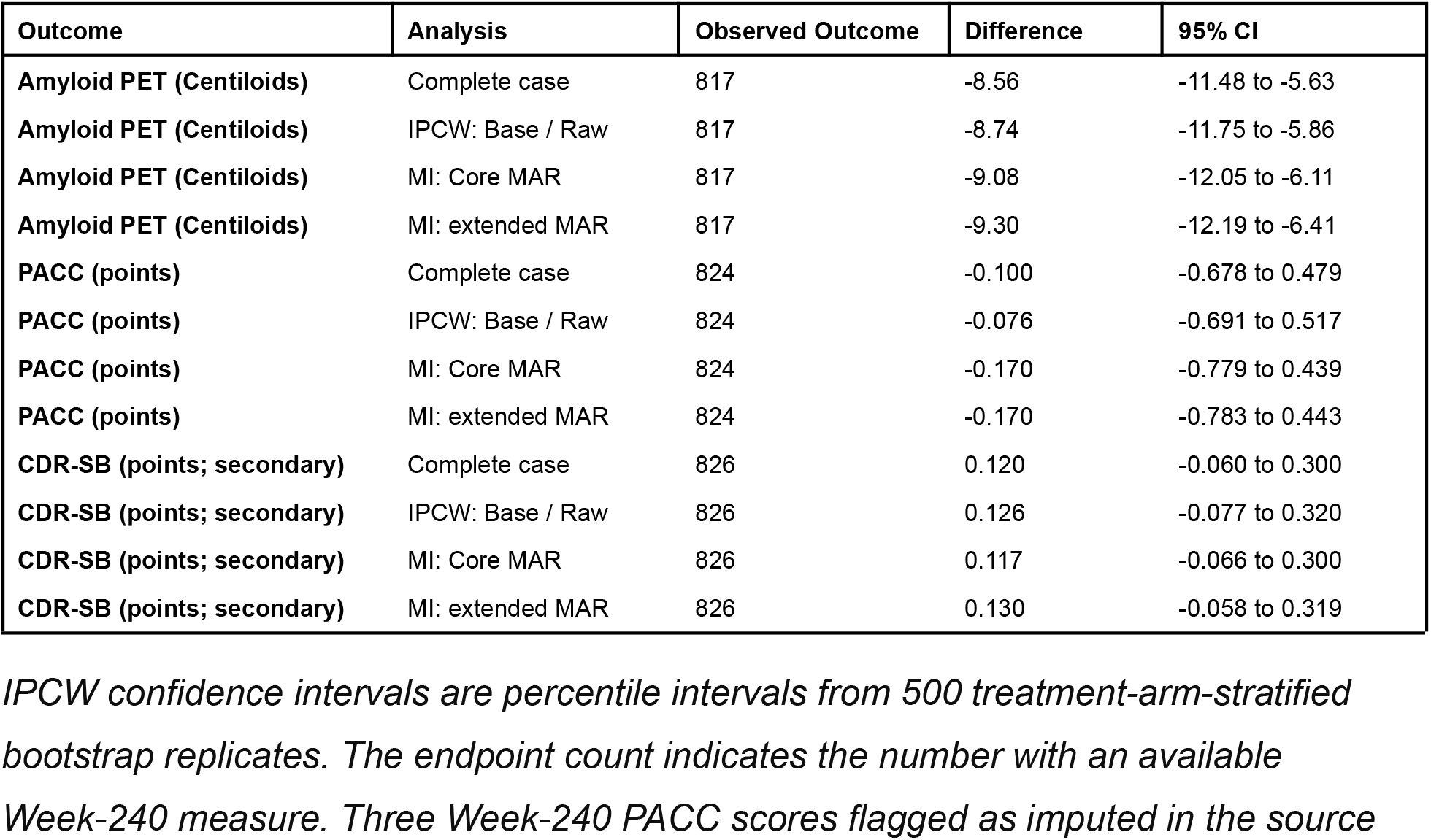
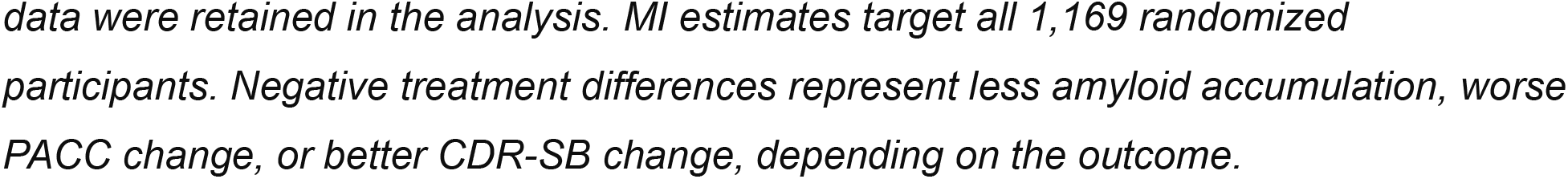
Treatment-assignment differences in baseline-to-week 240 change

### Multiple Imputation Under MAR

Core MAR imputation yielded treatment differences of -9.08 Centiloids (95% CI, -12.05 to -6.11), -0.170 PACC points (-0.779 to 0.439), and 0.117 CDR-SB points (-0.066 to 0.300). With the extended model, the differences were -9.30 Centiloids (-12.19 to -6.41), -0.170 PACC points (-0.783 to 0.443), and 0.130 CDR-SB points (-0.058 to 0.319; Table 2 and Figure 1). Monte Carlo error relative to standard error ranged from 7.1% to 7.7%. The amyloid difference remained negative, while neither cognitive nor functional outcome showed a clear treatment benefit under either MAR model.

### MNAR Sensitivity Analyses

The amyloid difference remained negative, with CIs excluding zero, across all six examined MNAR scenarios (Figure 2). When missing solanezumab outcomes were assumed to be 0.50 SD worse than MAR predictions, the difference were -3.48 Centiloids (95% CI, -6.50 to -0.46), -0.981 PACC points (95% CI, -1.602 to -0.360) and 0.297 CDR-SB points (95% CI, 0.112 to 0.481). A smaller 0.25 SD shift in the solanezumab arm yielded a CDR-SB difference of 0.207 points (95% CI, 0.024 to 0.390), whereas the PACC CI still included zero (-0.575; 95% CI, -1.188 to 0.038). Equal shifts in both arms changed the cognitive and functional differences little. In the tipping-point analysis, the amyloid CI first included zero when missing solanezumab outcomes were assumed to be 0.55 SD (19.7 Centiloids) worse than MAR predictions, whereas the PACC and CDR-SB CIs first excluded zero in the unfavorable direction at 0.30 SD (1.59 points) and 0.20 SD (0.24 points), respectively (Supplementary Figure S1).

**Figure 2.**
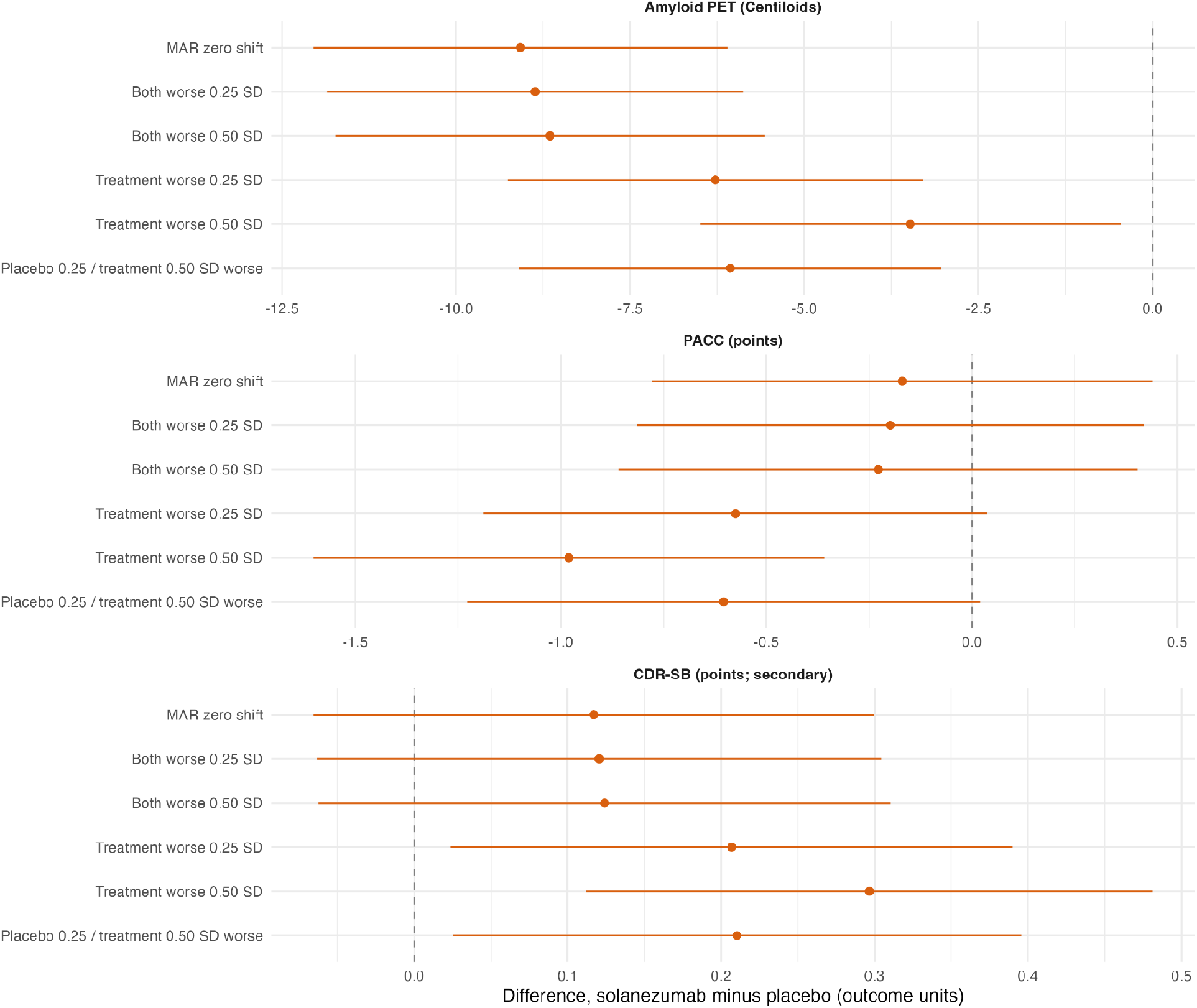
MNAR sensitivity of treatment-effect estimates. Panels display amyloid, PACC, and CDR-SB in their original units. Six scenarios shift only the missing Week-240 outcomes imputed under core MAR. Shifts are defined as 0.25 or 0.50 times the observed placebo-group Week-240 endpoint SD, with lower PACC and higher amyloid or CDR-SB considered worse. Observed outcomes are unchanged. Confidence intervals are conditional on the selected shifts.

## Discussion

Accounting for missing Week-240 outcomes had little effect on the main A4 treatment comparisons. Compared with the available-case analysis, IPCW changed the amyloid estimate by -0.18 Centiloids and the PACC estimate by 0.024 points. MAR imputation results were also close to those from the available-case and IPCW analyses. Consistent with prior trial results, across all approaches, solanezumab was associated with less amyloid accumulation, but no clear improvement in cognitive or functional outcomes.

Missing outcomes always reduce precision, but they bias treatment comparisons only when missingness is related to the unobserved outcome. IPCW and MAR imputation remove such bias only to the extent that measured variables explain it.

Our MNAR analyses therefore asked how far that assumption could fail before conclusions changed. The amyloid CI excluded zero until missing solanezumab outcomes were assumed to be about 20 Centiloids (0.55 SD) worse than MAR predictions, roughly twice the estimated effect, whereas PACC and CDR-SB showed unfavorable contrasts under smaller differential shifts (0.30 SD, 1.59 PACC points; 0.20 SD, 0.24 CDR-SB points), which would not be implausible if participants who left the solanezumab arm declined somewhat faster than predicted. MAR-based methods thus appear adequate for the amyloid result but do not settle the cognitive and functional results. These unfavorable contrasts reflect shifts imposed on missing outcomes, not evidence of harm.

### Strengths and Limitations

Strengths include analyzing the full randomized cohort, assessing missingness separately for each endpoint, and comparing approaches with different assumptions. However, IPCW and MAR imputation depend on the observed data adequately explaining missingness. Stable weights and consistent results across models cannot rule out bias from unmeasured factors. Some baseline predictors were filled with single values, and the MNAR shifts were chosen for illustration rather than estimated from the trial. Three Week-240 PACC composites in the A4 data package included an imputed component, introducing uncertainty not captured separately here. Disposition records could not establish whether missing assessments depended on unobserved cognitive status. For participants who died before Week 240, the Week-240 outcome may be undefined rather than missing, raising a separate estimand question. Finally, fixed-visit change does not use the full longitudinal trajectory, and A4’s cognitively unimpaired population and solanezumab exposure limit generalizability to trials of newer antibodies.

### Implications and Future Directions

In A4, adjustments based on measured baseline information left the main treatment-effect pattern largely unchanged, while the cognitive and functional outcomes remained sensitive to modest departures from MAR. Investigators of ongoing and future anti-amyloid trials, as well as those reanalyzing completed trials, should document why end-of-study outcomes are unavailable, distinguish missed assessments from treatment discontinuation or death, and report clinically plausible MNAR scenarios alongside MAR-based methods.

## Research in Context

**Systematic review:** Published A4 studies evaluated solanezumab’s efficacy and the potential of amyloid change as a surrogate for cognitive decline. These reports did not examine the sensitivity of Week-240 treatment differences to missing-outcome assumptions.

**Interpretation:** Methods that predict missing outcomes from measured information gave results nearly identical to analyses of participants with complete data. The amyloid result held even if participants with missing data were assumed to be much worse off, but the cognitive and functional results became unfavorable to solanezumab if those in the treatment arm were assumed to decline only modestly more than predicted.

**Future directions:** Investigators of ongoing and future anti-amyloid trials and those reanalyzing completed trials, should record why end-of-study outcomes are missing and report how conclusions change under plausible assumptions about participants with missing outcomes, particularly in populations with more cognitive decline and for drugs producing greater amyloid removal.

## Data Availability

Data is available to qualified investigators on a4studydata.org

http://a4studydata.org

## Acknowledgments

The A4 Study was a secondary prevention trial in preclinical Alzheimer’s disease, aiming to slow cognitive decline associated with brain amyloid accumulation in clinically normal older individuals. The A4 Study was funded by a public-private-philanthropic partnership, including funding from the National Institutes of Health-National Institute on Aging, Eli Lilly and Company, Alzheimer’s Association, Accelerating Medicines Partnership, GHR Foundation, an anonymous foundation, and additional private donors, with in-kind support from Avid Radiopharmaceuticals, Cogstate, Albert Einstein College of Medicine and the Foundation for Neurologic Diseases. The companion observational Longitudinal Evaluation of Amyloid Risk and Neurodegeneration (LEARN) Study was funded by the Alzheimer’s Association and GHR Foundation. The A4 and LEARN Studies were led by Dr. Reisa Sperling at Brigham and Women’s Hospital, Harvard Medical School, and Dr. Paul Aisen at the Alzheimer’s Therapeutic Research Institute (ATRI) at the University of Southern California. The A4 and LEARN Studies were coordinated by ATRI at the University of Southern California, and the data are made available under the auspices of Alzheimer’s Clinical Trial Consortium through the Global Research & Imaging Platform (GRIP). The complete A4 Study Team list is available on: https://www.actcinfo.org/a4-study-team-lists/. We would like to acknowledge the dedication of the study participants and their study partners who made the A4 and LEARN Studies possible.

GenAI Disclosure: Claude Opus 5.5 and ChatGPT-4 models were used for code debugging and minor language editing to improve concision to meet the word limit. All outputs were reviewed by multiple authors, who take full responsibility for the final manuscript.

Disclosure: SFA reports that Brown University received funding from Sanofi on her behalf for an unrelated student fellowship. She has received personal consulting fees from Boston University for work related to P01AG082653 and from Dayglen, LLC. She is the founder and a board member of the Center for the Future of Knowledge, a Rhode Island nonprofit corporation, and receives no compensation for these roles.

---

**Figure S1.**
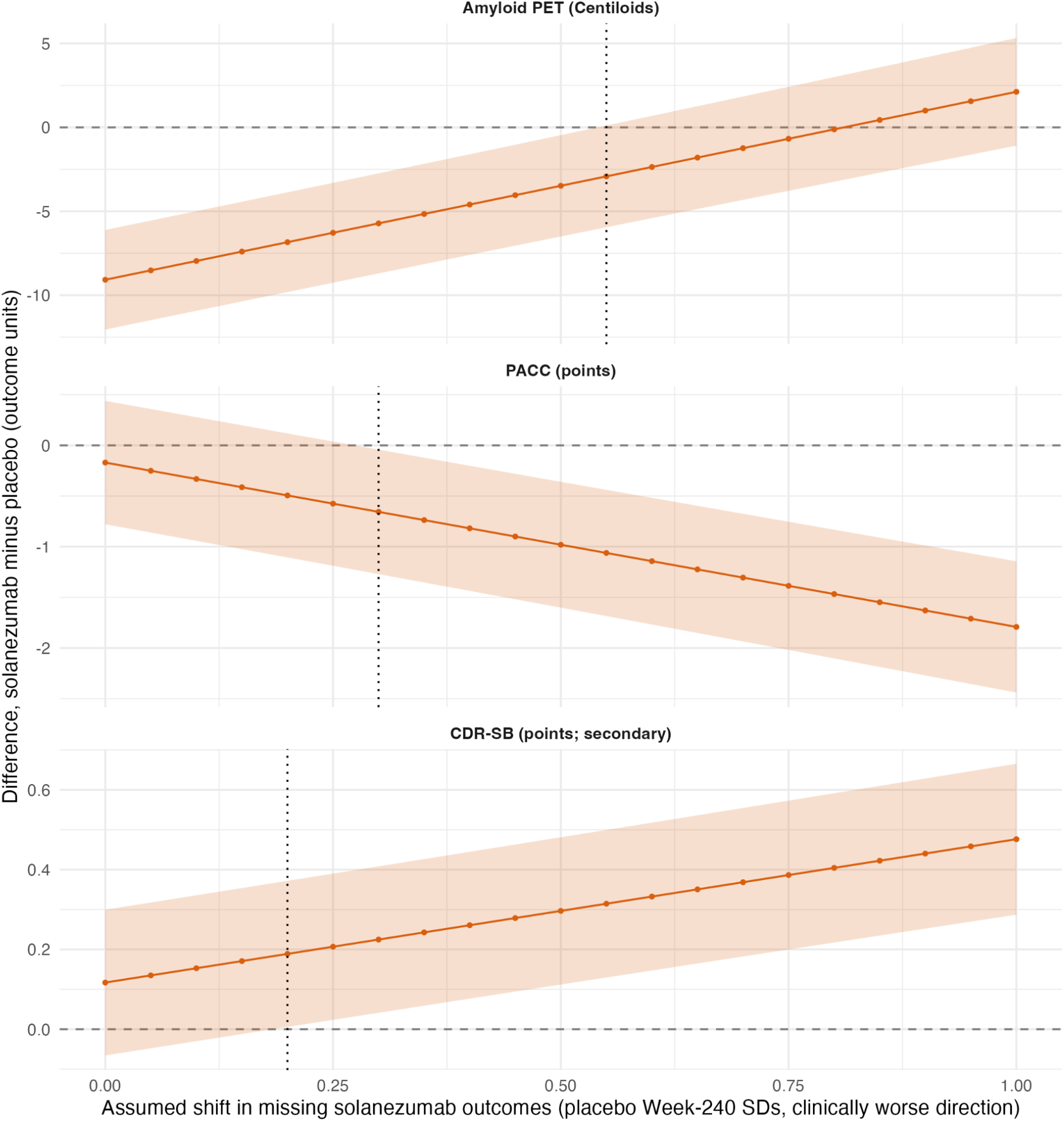
Tipping-point analysis. Treatment differences (solanezumab minus placebo) and 95% CIs as the assumed shift in missing solanezumab Week-240 outcomes increases from 0 to 1 placebo-arm SD (0.05 steps) in the clinically worse direction, with no shift in the placebo arm. Dotted lines mark the first shift at which the CI crosses zero (amyloid 0.55 SD, PACC 0.30 SD, CDR-SB 0.20 SD).

**Figure S2.**
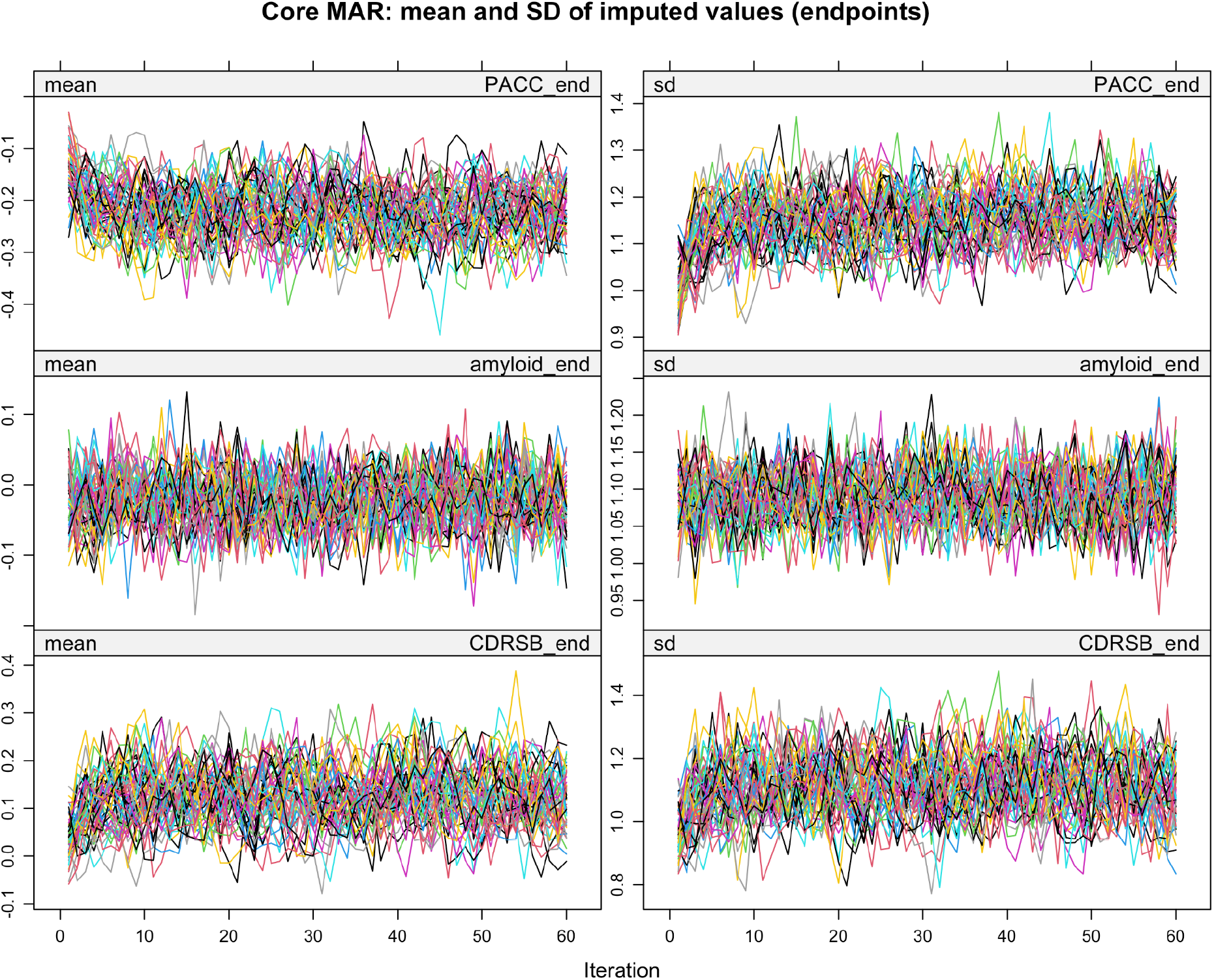
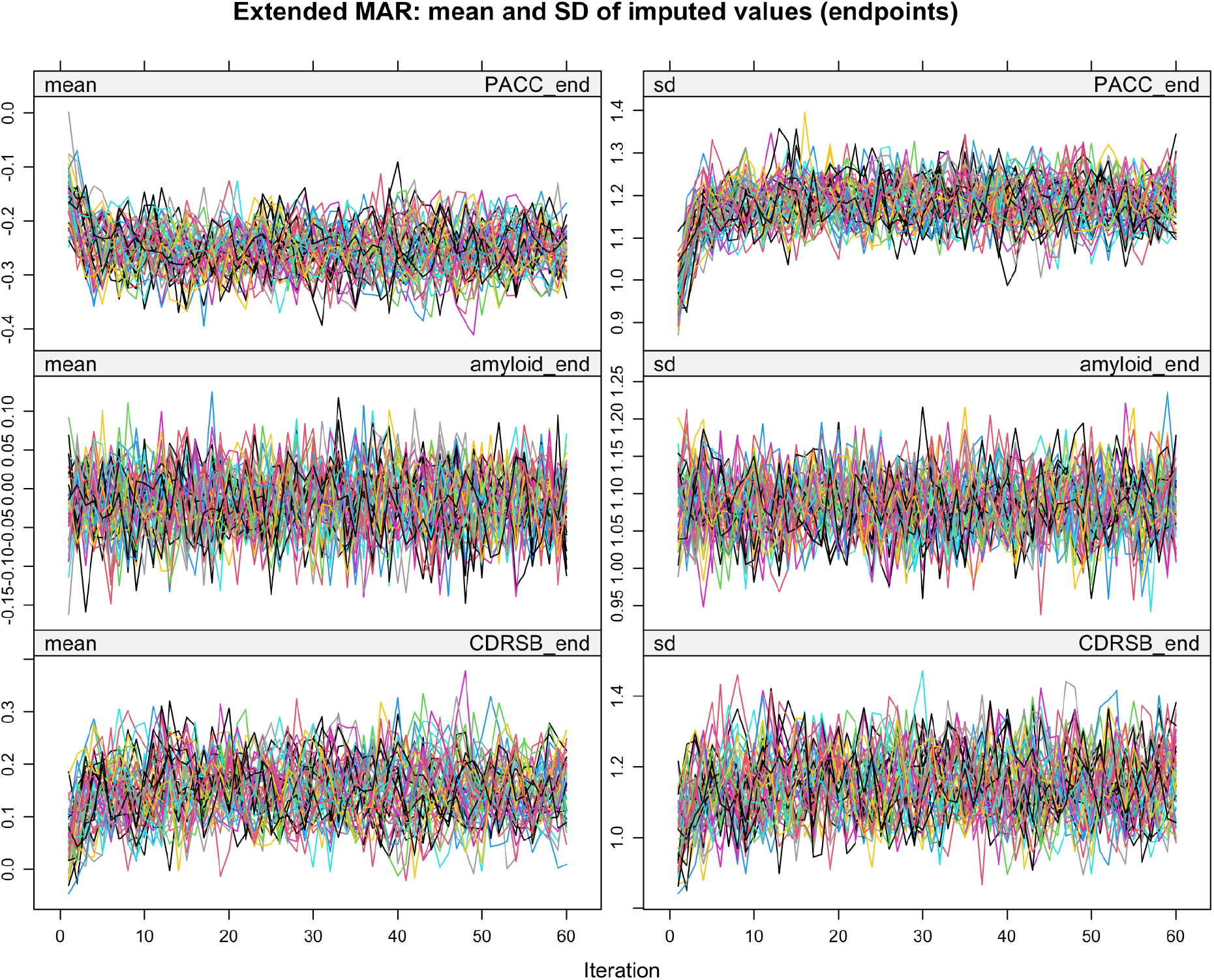
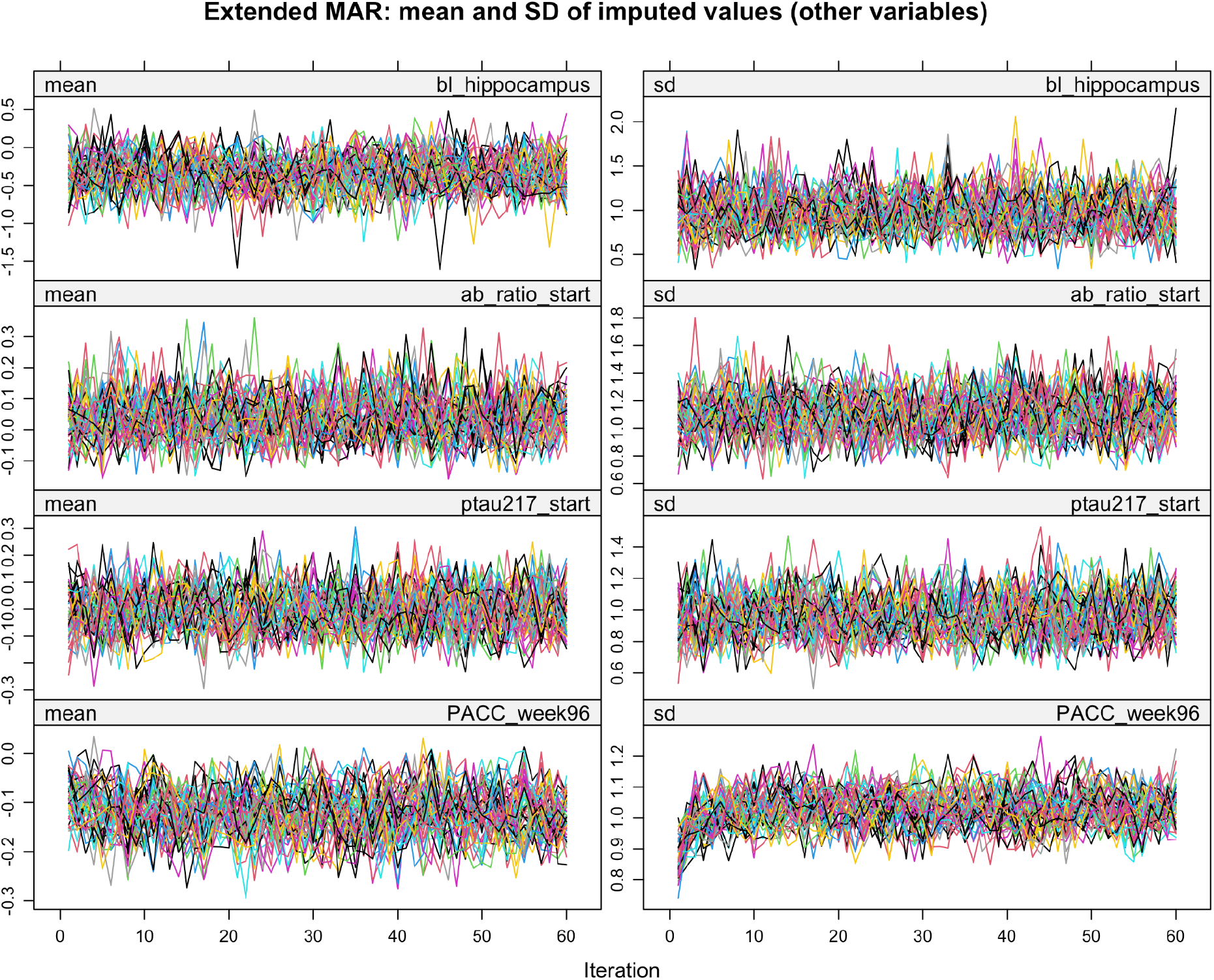
**MICE trace plots**. Mean and standard deviation of imputed values across 60 iterations for each of 50 imputation chains, for the core and extended MAR models. Numeric variables were imputed on the standardized scale, so values are in SD units. Variables with fewer than 10 missing values (BMI, n=2; microbleeds, n=1) are not shown.

**Table S1.** Variables included in each missing-outcome model. Every IPCW observation model used only pre-randomization variables. Both MAR models included the three Week-240 outcomes, while the extended model additionally included Week-96 PACC and other baseline predictors.

| **Model** | **Variables** | **Role** |
|---|---|---|
| IPCW: Base (primary) | Treatment assignment; age; sex; APOE ε4 copies (0, 1, 2); baseline amyloid PET (Centiloids); baseline PACC | Observation model, fit separately for each week 240 outcome |
| IPCW: + baseline microbleeds | Base + any definite or possible microbleed | Sensitivity |
| IPCW: + baseline characteristics | Base + education, years; BMI; Geriatric Depression Scale > 5 | Sensitivity |
| IPCW: + MRI volumes | Base + intracranial volume; forebrain parenchyma; hippocampus, entorhinal cortex, and lateral ventricle (left + right); hippocampal occupancy | Sensitivity |
| IPCW: + blood biomarkers | Base + plasma Aβ42/40; p-tau217; GFAP; NfL | Sensitivity |
| IPCW: + baseline cognition | Base + MMSE; Logical Memory delayed recall; FCSRT total recall | Sensitivity |
| IPCW: + all baseline predictors | Base + all variables above | Sensitivity |
| MI: Core | Treatment assignment; age; sex; APOE ε4 copies; baseline and Week-240 amyloid PET, PACC and CDR-SB | MAR imputation; basis for MNAR scenarios and tipping-point analyses |
| MI: Extended | Core + education; BMI; Geriatric Depression Scale > 5; hippocampal volume; any microbleed; plasma Aβ42/40 and p-tau217; MMSE and DSST; PACC at Week 96 | MAR imputation |

**Table S2.** Visit and missing values for each model variable. Missing values among baseline variables ranged from none for the design and demographic variables to 57.6% for plasma NfL. Missing GFAP and NfL values almost entirely reflected participants without a recorded result (499 participants had GFAP and NfL records); only three NfL results were set to missing because they were below the quantification limit or had insufficient sample. All baseline variables were collected before randomization.

| Variable | Visit | Missing, n (%) | Model |
|---|---|---|---|
| **Study design and demographics** |  |  |  |
| Treatment assignment | Randomization | 0 | IPCW, MI |
| Age, years | Screening | 0 | IPCW, MI |
| Sex | Screening | 0 | IPCW, MI |
| APOE ε4 copies | Screening | 0 | IPCW, MI |
| Education, years | Screening | 0 | IPCW, MI |
| BMI | Screening | 2 (0.2) | IPCW, MI |
| Geriatric Depression Scale >5 | Screening | 0 | IPCW, MI |
| **Baseline outcome measures** |  |  |  |
| CDR-SB | Visit 1 | 0 | MI |
| Amyloid PET, Centiloids | Visit 2 | 0 | IPCW, MI |
| PACC | Visit 6 | 0 | IPCW, MI |
| **Baseline cognition** |  |  |  |
| MMSE | Visit 6 | 0 | IPCW, MI |
| Logical Memory delayed recall | Visit 6 | 5 (0.4) | IPCW |
| FCSRT total recall | Visit 6 | 8 (0.7) | IPCW |
| DSST | Visit 6 | 0 | MI |
| **Baseline MRI** |  |  |  |
| Intracranial volume, cm³ | Visit 4 | 10 (0.9) | IPCW |
| Forebrain parenchyma volume, cm³ | Visit 4 | 10 (0.9) | IPCW |
| Hippocampal volume (left + right), cm³ | Visit 4 | 10 (0.9) | IPCW, MI |
| Entorhinal volume(left + right, cm³ | Visit 4 | 10 (0.9) | IPCW |
| Lateral ventricle volume (left + right), cm³ | Visit 4 | 10 (0.9) | IPCW |
| Hippocampal occupancy | Visit 4 | 10 (0.9) | IPCW |
| Any microbleed (definite or possible) | Visit 4 | 1 (0.1) | IPCW, MI |
| **Baseline plasma biomarkers** |  |  |  |
| GFAP | Visit 1 | 670 (57.3) | IPCW |
| NfL | Visit 1 | 673 (57.6) | IPCW |
| Aβ42/40 ratio | Visit 3 | 350 (29.9) | IPCW, MI |
| p-tau217 | Visit 6 | 87 (7.4) | IPCW, MI |
| **Post-randomization measures (MI only)** |  |  |  |
| PACC, Week 96 | Visit 30 | 180 (15.4) | MI |
| PACC, Week 240 | Visit 66 | 345 (29.5) | MI (outcome) |
| Amyloid PET, Week 240 | Visit 66 | 352 (30.1) | MI (outcome) |
| CDR-SB, Week 240 | Visit 66 | 343 (29.3) | MI (outcome) |

## Notes

### Clinical Trial

NCT02008357

### Author Declarations

This work represents a secondary data analysis using deidentified individual-level data from the A4 study. All study participants provided informed consent at the time of enrollment in the study. IRB approval was obtained at each of the trial sites. The use of deidentified data in the present analysis was determined to be exempt from additional IRB review via consultation with the Brown University IRB staff.

